# Early experiences of rehabilitation for patients post-COVID to improve fatigue, breathlessness exercise capacity and cognition

**DOI:** 10.1101/2021.03.25.21254293

**Authors:** E Daynes, C Gerlis, E Chaplin, N Gardiner, S Singh

**Affiliations:** Centre of Exercise and Rehabilitation Sciences, NIHR Leicester Biomedical Research Centre-Respiratory, University Hospitals of Leicester, UK; Department of Respiratory Sciences, University of Leicester, UK

**Author notes:** Conflicts of interest: nil to disclose.

## Abstract

Patients with lasting symptoms of COVID-19 should be offered a comprehensive recovery programme. Patients that completed a six week, twice supervised adapted pulmonary rehabilitation programme demonstrated statistically significant improvements in exercise capacity, respiratory symptoms, fatigue and cognition. Participants improved by 112m on the Incremental Shuttle Walking Test and 544 seconds on the Endurance Shuttle Walking Test. There were no serious adverse events recorded, and there were no dropouts related to symptom worsening. COVID-19 rehabilitation appears feasible and significantly improves clinical outcomes.

## Introduction

COVID-19 can lead to a number of lasting symptoms such as breathlessness, fatigue and reduced ability to engage in activities of daily living^1^. It became apparent that a recovery or rehabilitation programme would be necessary to support those to return to normal following infection. The European Respiratory Society taskforce identified a need for a formal assessment to understand physical and emotional functioning to determine rehabilitation needs following COVID-19 infection^2^. This statement identifies the need for a rehabilitative intervention following hospital discharge and includes a comprehensive programme. It has been acknowledged that a unidimensional programme will not meet the needs of COVID-19 survivors who will present with a variety of different symptoms^3^. However, there is invariably some overlap between the needs of COVID-19 survivors and a Pulmonary Rehabilitation (PR) population. PR is a highly evidenced based intervention and addresses many symptoms reported in the post COVID population, but it will likely need modifications. Therefore this model was adapted when developing COVID-19 rehabilitation to meet the complex needs of these patients^2^. This study presents the results of the initial COVID-19 rehabilitation programme using a modified PR programme.

## Methods

This study reports the experiences of the first 32 patients that completed rehabilitation following COVID-19 infection. This observational study was approved by the National Health Service Research Ethics Committee (reference 17/EM/0156) and registered through the ISRCTN (ISRCTN45695543). Patients were referred through a discharge follow up pathway; at COVID-19 medical follow up or; a referral from their GP. Patients were included if they self-identified rehabilitation needs, and were excluded if they demonstrated acute symptoms or were not medically stable. Patients that had COVID-19 but managed in the community were eligible for the programme and were referred by their GP. All patients were screened for unexplained symptoms and unstable cardiovascular disease.

The rehabilitation programme was six weeks in duration, with two supervised sessions per week. The programme comprised of aerobic exercise (walking/treadmill based), strength training of upper and lower limbs and educational discussions with handouts from the www.yourcovidrecovery.nhs.uk website. The Borg breathlessness scale and rate of perceived exertion were used alongside self-reported symptoms to determine progression of the exercises. The outcomes were: the incremental and endurance shuttle walking test (ISWT/ESWT), COPD Assessment Test (CAT)^4^, Functional Assessment of Chronic Illness Therapy Fatigue Scale (FACIT), Hospital Anxiety and Depression Scale (HADS), EuroQual 5 domains (EQ5D) and the Montreal Cognitive Assessment (MoCA). The ISWT and ESWT were completed in line with gold standards on a 10m course and performed a familiarisation test at baseline^5^.

Data was analysed using SPSS v25. Patients were considered completers if they attended eight out of 12 scheduled sessions. A paired t-test was used to compare changes before and after rehabilitation.

## Results

30 patients completed the COVID-19 rehabilitation programme (52% male, mean [SD] age 58[16] years, length of stay 10[14] days). 5(14%) patients required mechanical ventilation during their admission and were managed in an ITU setting. The mean[SD] time from confirmed infection to enrolment onto the programme was 125[54] days. The mean [SD] number of sessions completed was 11[2] out of a scheduled 12 sessions. 30 patients completed at least eight sessions of rehabilitation, with 2 drop outs due to social circumstances.

Baseline scores identified reduced exercise capacity and health related quality of life but relatively well preserved anxiety, depression and cognition as measured by the HADS and MoCA respectively. Patients that completed rehabilitation had a mean [SD] improvement in the ISWT of 112[105]m (p<0.01), and 544[377] seconds (p<0.01). The FACIT improved by 5[7] points (p<0.01), the EQ5D thermometer improved by 8[19] (p=0.05) and MoCA by 2[2] points (p<0.01) The CAT score improved by a score of 3[6] (p<0.05).The HADS anxiety and depression scores improved by 0[4] and 1[4] respectively which was not statistically significant, however the baseline scores were low (table 1). Figure 1 demonstrates the changes in FACIT and ISWT.

**Table 1:** Clinical outcomes pre and post COVID rehabilitation.

| N=30 | Pre rehabilitation | Post rehabilitation | Change | P= |
| --- | --- | --- | --- | --- |
| ISWT (m) | 300[198] | 413[229] | 112[105] | <0.01 |
| ESWT (seconds) | 292[260] | 837[406] | 544[377] | <0.01 |
| CAT | 16[7] | 13[7] | -3[6] | <0.05 |
| FACIT | 29[14] | 34[13] | 5[7] | <0.01 |
| EQ5D Thermometer | 62[18] | 70[21] | 8[19] | 0.05 |
| MoCA | 25 [3] | 27[3] | 2[2] | <0.01 |
| HADS A | 6[4] | 6[5] | 0[4] | 0.5 |
| HADS D | 6[4] | 5[4] | -1[4] | 0.1 |

**Figure 1.**
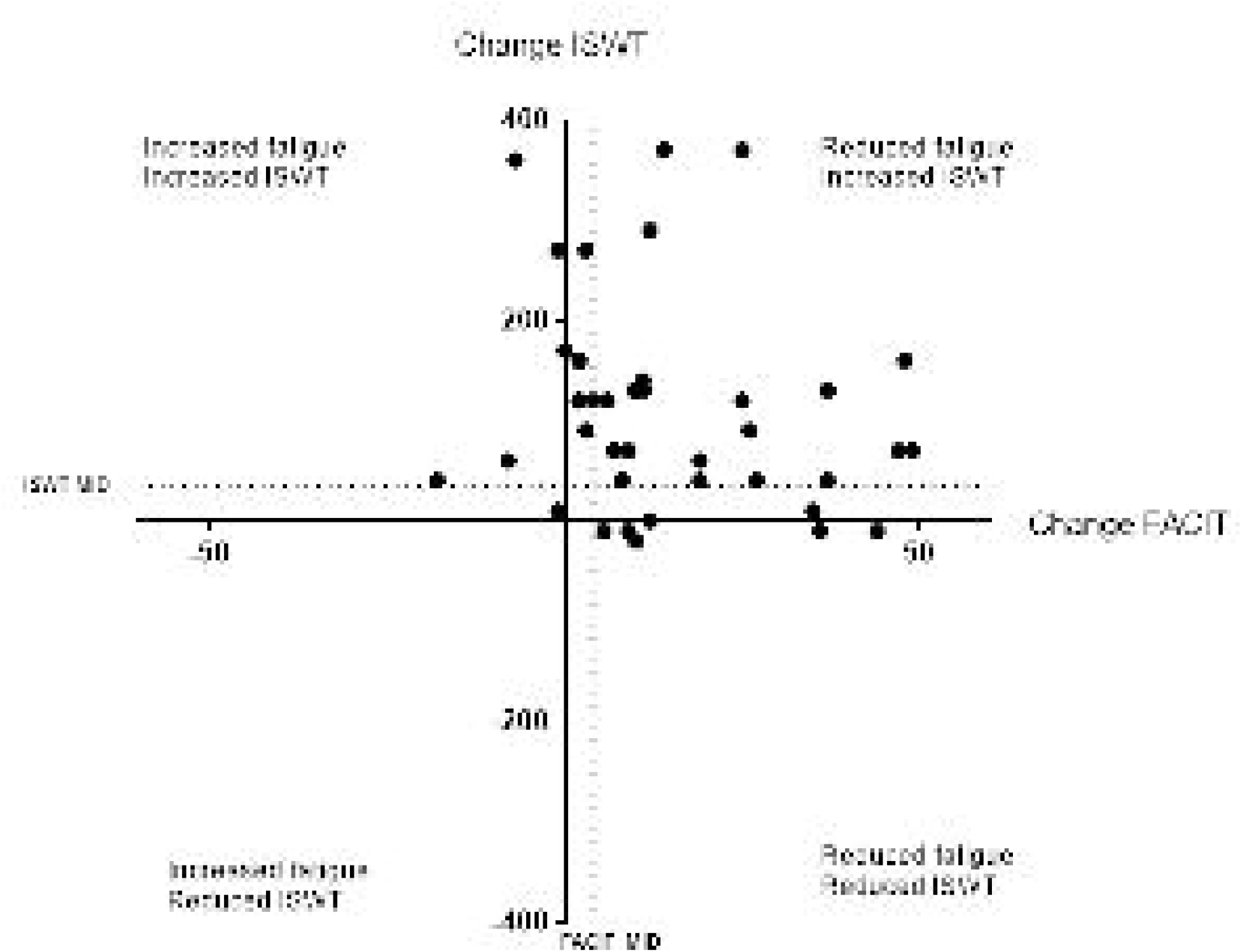
Changes in Fatigue and Incremental Shuttle Walking Test for patients completing a COVID-19 Rehabilitation programme.

Table 1 Mean[SD] of clinical outcomes pre and post COVID rehabilitation. ISWT Incremental Shuttle Walking Test, ESWT Endurance Shuttle Walking Test, CAT COPD Assessment Test, FACIT Functional Assessment of Chronic Illness Therapy (fatigue score), EQ5D EuroQual 5 Domain, MoCA Montreal Cognitive Assessment, HADS Hospital Anxiety and Depression Scale (A Anxiety and D Depression domain).

## Discussion

This adapted rehabilitation programme for patients following COVID-19 has demonstrated feasibility and promising improvements in clinical outcomes. There was a high completion rate of COVID rehabilitation in the first 32 patients as demonstrated by this study, with no dropouts attributed to a worsening of symptoms, indicating acceptability to the participants of the intervention. There were significant improvements in clinical outcomes of walking capacity and symptoms of fatigue, cognition and respiratory symptoms (measured by the CAT). Whilst there are no known minimal important differences for these outcomes in patients with COVID-19; the ISWT, ESWT, CAT, FACIT and, MoCA all exceed the known minimal important differences in patients undergoing conventional pulmonary rehabilitation, or with a chronic respiratory condition. It is possible that there is some natural recovery in this cohort of patients, however, as the mean length of time between infection and enrolment onto the programme was 125 days it is likely that natural recovery had slowed down.

There has been concern that rehabilitation may worsen or trigger symptoms of post-viral fatigue and that exercise therapy may exacerbate fatigue. The exercise element of this programme is progressed (by staff experienced in delivering pulmonary and cardiac rehabilitation programmes) in line with patient’s symptoms throughout the programme and provides a holistic and pragmatic approach to exercise therapy. The educational component of the programme supports management techniques useful for these symptoms (such as pacing and prioritising). There were no serious adverse events during the course of the programme supporting the safety of this intervention. The majority of patients improved both the symptom of fatigue and exercise capacity. One patient did not improve either fatigue or exercise capacity (due to a previous stroke). The remainder (n= 4) who reported an increase in fatigue recorded meaningful improvements in their exercise capacity.COVID-19

Rehabilitation programmes should aim to provide a holistic and multi-faceted approach to managing post-COVID symptoms. Clinicians should aim to individualise programmes and to monitor adverse events and symptoms, given the limited evidence in the field.

To conclude, an adapted pulmonary rehabilitation programme to support patients with lasting symptoms of COVID-19 is safe and demonstrates improvements in exercise capacity and symptoms of breathlessness, fatigue and cognition.

## Data Availability

The authors confirm that the data supporting the findings of this study are available within the article [and/or] its supplementary materials.

## References

1. Cortinovis, N., Perico, N., Remuzzi, G. Long-term follow-up of recovered patients with COVID-19 The Lancet 2021:397(10270): 173–175.

2. Spruit, M., Holland, A., Singh, S., et al. COVID-19: Interim Guidance on Rehabilitation in the Hospital and Post-Hospital Phase from a European Respiratory Society and American Thoracic Society-coordination International Task Force. ERJ 2020 57(1).

3. British Thoracic Society. Delivering rehabilitation to patients surviving COVID-19 using an adapted pulmonary rehabilitation approach – BTS guidance. [Online] Available from www.brit-thoracic.org.uk (2020) [Accessed on 08/02/2021)

4. Daynes, E., Gerlis, C., Briggs-Price, S., et al. COPD assessment test for the evaluation of COVID-19 symptoms. Thorax 2020 0(1-3).

5. Holland, A., Spruit, M., Troosters, T., et al. An Official European Respiratory Society/American Thoracic Society technical standard: field walking tests in chronic respiratory disease. ERJ (2014): 44:1428–1446

